# Protocol for VIVALDI Social Care: Pilot Study to reduce Infections, Outbreaks and Antimicrobial Resistance in Care Homes for Older Adults

**DOI:** 10.1101/2023.11.20.23298504

**Authors:** M Krutikov, Z Fry, B Azmi, C Lezard, K Thorn, G Patefield, G Childe, J Hudson, O Stirrup, A Jhass, N Turner, J Cassell, P Flowers, A Hayward, A Copas, M Green, L Shallcross

## Abstract

Care home residents are vulnerable to severe outcomes from infections such as COVID-19 and influenza. However, measures to control outbreaks, such as care home closures to visitors and new admissions, have a detrimental impact on their quality of life. Many infections and outbreaks could be prevented but the first step is to measure them reliably. This is challenging in care homes due to the lack of data and research infrastructure. During the pandemic, the VIVALDI study measured COVID-19 infections in residents and staff by partnering with care providers and using routinely collected data. This study aims to establish sentinel surveillance and a research database to enable observational and future interventional studies in care homes. The project has been co-produced with care providers, staff, residents, relatives, and researchers.

The study (October 2023 to March 2025) will explore the feasibility of establishing a network of 500-1500 care homes for older adults in England that is underpinned by a linked data platform. No data will be collected from staff. The cohort will be created by regularly extracting resident identifiers from Digital Social Care Records (DSCR), followed by pseudonymisation and linkage to routinely collected datasets.

Following extensive consultation, we decided not to seek informed consent from residents for data collection, but they can ‘opt out’ of the study. Our goal is to be inclusive, and it is challenging to give every resident the opportunity to ‘opt in’ due to cognitive impairment and the requirement for consultees. The project, and all requests to use the data will be overseen by relatives, residents, staff, and care providers. The study has been provisionally approved by the Health Research Authority Confidentiality Advisory Group and the South-West Frenchay Research Ethics Committee. It is funded by the UK Health Security Agency.

## Plain English Summary

Infections like flu or COVID-19 are common in care homes and infected residents can become seriously unwell. When infections spread, the measures that are commonly used to stop outbreaks, like care home closures to visitors and new admissions, can have a detrimental impact on residents because their family or friends cannot visit. The first step to solving this problem is being able to measure how often infections and outbreaks happen, and how this varies across care homes. This is currently difficult because there are no systems to collect data from care home residents.

During the COVID-19 pandemic, care homes worked with researchers and the Government to deliver a research study called VIVALDI which measured COVID-19 infections in residents and staff and monitored what happened to them. This pilot study builds on what we learned in the pandemic and aims to reduce the impact of common infections (like flu, diarrhoea and vomiting) on residents. In the study we will set up a network of 500-1500 care homes for older adults in England that are interested in research. By collecting limited data (NHS numbers) from residents in these homes and linking to other datasets that are already held in the secure NHS environment, we will be able to measure the extent of common infections in residents. We are not collecting data from staff, and it will not be possible to identify any residents in the datasets. We will also create an anonymous database (names, dates of birth, NHS numbers removed), that can be used by researchers to find new ways to prevent infection in care homes. This dataset will be stored securely by the research team at UCL. If the project is successful, and residents and relatives support continued use of their data in this way, we hope that this approach will be used to monitor infections in care homes on a permanent basis.

The study has been designed in partnership with care providers, experienced care staff, policymakers, residents and their relatives, and academics. Relatives, residents (where possible), care home staff and care providers will oversee the study and all research outputs.

## Introduction

In England, around 380,000 people live in 11,000 care homes for older adults and this number is increasing.^1^ The majority of care home residents are older than 85 years, at least two-thirds live with dementia, and average life expectancy within the care home is 12 to 24 months.^2,3^ The COVID-19 pandemic has highlighted the extreme vulnerability of residents to severe outcomes following infection.^4^ However, every year Antimicrobial Resistance (AMR), respiratory (e.g., influenza, pneumonia), gastrointestinal (e.g., norovirus), urinary tract and skin infections cause substantial morbidity and mortality and cause outbreaks which have negative consequences like closure of care homes to visitors and new admissions, and avoidable hospital admissions.^5,6^ Many of these infections and outbreaks could be averted by implementing effective interventions, and generating new evidence on how to prevent or reduce infection where none currently exists.

Efforts to reduce the impact of infection are hampered by multiple, complex barriers including an inadequate research and data infrastructure, the fragmentation of social care, multiplicity of providers, and poor integration of health and social care and other services that impact on health and wellbeing. The sector has also failed to benefit from advances in care driven by National Institute for Health and Care Research (NIHR) which is primarily focused on hospitals and primary care. The urgent need to address this gap in care home specific data and surveillance, which severely undermined the initial response to COVID-19 in care homes, was recently highlighted by the Chief Medical Officer in his Technical report on the COVID-19 pandemic in the UK.^7^

Prior to the pandemic, the need for better quality individual-level data from care homes to enable comparison of care quality and outcomes was widely acknowledged, however progress towards this goal was limited.^8^ The NIHR-funded “Developing research resources and minimum dataset for care homes adoption and use” (DACHA) study was established to help address this research gap, by synthesising existing evidence and data sources with care home generated resident data, to deliver an agreed minimum data set that is usable and authoritative for different user groups.^9^ However the urgency of the COVID-19 pandemic catalysed new, complementary initiatives to improve the availability of data from care homes, including the UCL-led and Government-funded VIVALDI observational study [<u>VIVALDI Study</u> <u>| UCL Institute of Health Informatics - UCL – University College London</u>], which ran from 29th May 2020 until 31st March 2023.^10^ VIVALDI measured infections, immunity, and vaccine effectiveness in care home staff and residents over successive waves of the pandemic.^11^ Study findings enabled policymakers to make difficult but evidence-based decisions to protect the sector and demonstrated how timely surveillance and research on both can inform policy and reduce the impact of infections. It also showed that it is feasible to deliver research and surveillance on infection at scale and pace in care homes. Key to VIVALDI’s success was the establishment of strong partnerships with providers, use and linkage of routinely collected data, and close collaboration between academics and policymakers. Importantly, the care sector has been fully engaged in this process which has demonstrated how research can directly benefit them.

The first step in being able to deliver research on infection, outbreaks and AMR in care homes is to quantify the burden and outcomes of different infections in residents at scale. Care homes cannot collect this information manually; therefore, the best approach is to access data that are already being collected by the National Health Service (NHS) and public health systems. However, identifying residents within routine data is challenging. The VIVALDI study overcame this problem by capitalising on the fact that care home residents were regularly being tested for COVID-19.^12^ This created an accurate ‘registry’ of residents linked to their care home, which could be linked to other routinely collected datasets via residents’ NHS numbers, a unique identifier. Now COVID-19 testing has stopped,^13^ this approach is no longer viable. Address-based matching has been applied in some research studies to identify residents; however, the sensitivity and specificity of this approach in care homes varies^4,14^ particularly given rapid changes to the resident population which limit the representativeness of any data generated. In addition, addresses that are recorded in NHS medical records are often inaccurate as they are time lagged, incomplete, and do not include temporary residents (receiving respite or step-down care); while data from local authorities, which commission some beds, does not include residents who are self-funded. Recent hospital admissions are also likely among temporary residents who may therefore import infection into the care home – a risk highlighted in the pandemic^15^ – so including them in surveillance and research studies is essential.

The study described in this protocol aims to measure infections and AMR in care homes (in collaboration with the UK Health Security Agency (UKHSA)), and to establish a database that can be used for observational research, and in future interventional studies, on infectious diseases, outbreaks, and AMR to inform policy and practice. We aim to include data from approximately 15,000-30,000 care home residents in England. Building on the experiences from the VIVALDI study, Digital Social Care Record (DSCR) suppliers will share NHS numbers belonging to residents within participating care homes which will enable identification of the study cohort and linkage across routine datasets to capture data on infections, vaccinations, and clinical outcomes (hospital admissions, deaths). The study protocol has been informed by extensive engagement with relatives or care home residents, care staff, providers, and visits to care homes to talk to residents which have confirmed that reducing the impact of common infections and outbreaks is a priority for people who live and work in this setting. Engagement activities have been enabled by a new partnership between the UCL research team, Care England, the largest care sector representative organisation, and the Outstanding Society (OS), a community interest company who promote quality in social care.

### Protocol

#### Study design

Research Database, Prospective Open Cohort Study

#### Settings

Care Quality Commission (CQC) registered care homes that provide nursing or residential care for older people (aged >=65 years) in England and use DSCR.

#### Participants

Residents in participating care homes

##### Inclusion criteria

Residents who have not opted out of data collection.

##### Exclusion criteria

Residents of care homes that primarily provide care to adults < 65 years, although residents who are younger than 65 years who reside in care homes that primarily provide care to older adults will be eligible for inclusion.

#### Aims & Objectives

Aim: To test the feasibility of establishing sentinel surveillance and a research database to facilitate large-scale observational research and future interventional studies to reduce infection, outbreaks, and AMR in care homes for older adults in England.

##### Primary objective

To establish a care home network, underpinned by linked data to enable sentinel surveillance and research on infection and AMR in older adult residents of 500-1500 care homes in England.

##### Secondary objectives

● To engage effectively with care home stakeholders to support effective design and delivery of the study.
● To recruit 500-1500 care homes to take part in the study.
● To establish information governance approvals and templates to enable sharing of data on residents.
● To establish data pipelines to extract individual-level data on care home residents and transfer these data securely to NHS England.
● To pseudonymise and link individual-level data on care home residents to routinely collected datasets held in a secure data platform, like the NHS Foundry, to create the research database.
● To establish a Data Access Committee to oversee future use of the research database and enable its use by researchers.
● To deliver an exemplar study on antimicrobial prescribing and resistance in care home residents using the database.
● To engage with the care sector and policymakers to disseminate study findings.

#### Outcomes

##### Primary outcomes

The establishment of sentinel surveillance and a research database which enables observational and future interventional studies on infection in older adult residents of care homes in England.

##### Secondary outcomes

● Study design and delivery that has been co-produced with care home stakeholders. Descriptions and reflections on the process will be published and disseminated.
● A network of 500-1500 participating care homes with appropriate contracts in place.
● Appropriate ethical and data governance approvals in place.
● Effective data pipelines for extraction of individual-level data on care homes residents, pseudonymisation, and linkage to routinely collected datasets to create a research database.
● An effective Data Access Committee comprising key stakeholders with established ways of working that can be implemented in future research.
● Effective bespoke data opt-out system for residents to opt-out of sharing that can be used for future databases.
● Incidence of AMR and modelling of association with antimicrobial prescribing practice in care home residents.

#### Consent

A key feature of the project is its goal to be inclusive, so every resident has the opportunity to take part, and findings are relevant for *all* older adults in care homes. It is also important that the database is accurate, up-to-date, and complete, for studies which aim to measure the burden of infection and related outcomes. This is because infections spread between people, and short-stay residents (who often get excluded from research and studies that are based on routinely collected data) are more likely to bring infections into care homes having recently been in hospital.

An estimated 70% of care home residents live with cognitive impairment^16^ and may not be able to provide informed consent to study participation. Whilst it is possible to identify consultees to support the consent process in those who lack capacity; this is time-consuming and requires dedicated staff. Severe staff shortages across the care sector and lack of embedded research infrastructure mean it is not feasible for staff to seek consent from every care home resident. Consequently, to develop an inclusive study and for data to be generalisable, the study has sought Section 251 support from Health Research Authority Confidentiality Advisory Group (HRA CAG) to collect data from *all* residents without consent, with the option for residents to opt-out of data sharing. In large care provider organisations, the initial decision to take part will be taken by their senior management team and home managers will then be asked to confirm if their care home is willing to take part. In homes that are not part of larger organisations, the decision to take part will be made by the care home manager.

#### Care home engagement and recruitment

Since October 2022, researchers from UCL have been working with Care England and the Outstanding Society to raise awareness of the VIVALDI Social Care Project through presentations at Care conferences and events held by the CQC, press releases, magazine articles, newsletters, webinars, our organisations’ websites and podcasts (https://theoutstandingsociety.co.uk/case-study/vivaldi-care-home-study/). As of May 2023, over 800 care providers in England have expressed an interest in taking part in the study.

When the study begins, the research team will distribute multi-modal study materials to care homes that provide information at three levels: 1) basic information about the study including how to opt out of data collection, 2) detailed information about the study, and 3) very detailed information including study design, explanation of the data flows and data controllership. These materials have been co-developed with the study’s *Engagement Working Group* which includes relatives, care home staff, and providers, and piloted with a subset of care homes prior to study start.

#### Data Collection & Management

Engagement work with the care sector has made it clear that it is not feasible to extract data from paper-based systems. Following extensive discussions with care providers and the Digital Care Hub (formerly Digital Social Care), a sector led organisation which provides free support to care providers on technology, data protection, and cyber security (www.digitalcarehub.co.uk), it has been agreed to start by working with providers that use DSCR that have been assured by the NHS Transformation Directorate in NHS England (NHSE).^17^

To maintain an accurate record of which residents are in a care home at any point in time, data for each resident will be extracted directly from DSCR rather than from the care homes. This will include residents’ NHS numbers, the date of data collection and the unique care home identifier allocated by the Care Quality Commission (CQC ID) at regular intervals (residents who have opted out of data collection will be excluded). This will be achieved by establishing a permanent daily data feed from the DSCR supplier to a secure environment within NHSE, allowing the dataset to be updated in near real-time. Although this may impact on the generalisability of the sample for the pilot study, the Government has strongly signalled its expectation that all care providers will move to electronic care records (80% of CQC registered providers to have DSCR by March 2024).^18^ Currently over 60% of providers use DSCR^19^ suggesting it will become increasingly feasible to recruit a diverse set of care homes that use DSCR over time, if the study is continued beyond the pilot. To reduce the administrative burden associated with participating, a set of template agreements (data sharing agreements, data privacy impact assessment, privacy notice) will be developed in partnership with Digital Care Hub and key stakeholders for use by participating providers.

The linked data platform will be established in a secure data environment within NHSE, such as NHS Foundry,^20^ with NHSE acting as data processors on behalf of the project. NHSE will first pseudonymise data from residents using an automated algorithm and then allocate each record a common pseudo-identifier, based on NHS number. This can link records from residents to routine collected datasets already held in this secure data environment (hospital admissions, vaccinations, emergency admissions, microbiology and virology results, antimicrobial prescribing, mortality), Figure 1, Table 1. Set-up and curation of the linked data platform in the secure data environment will be led by a software engineer who oversees the VIVALDI data platform.

**Figure 1:**
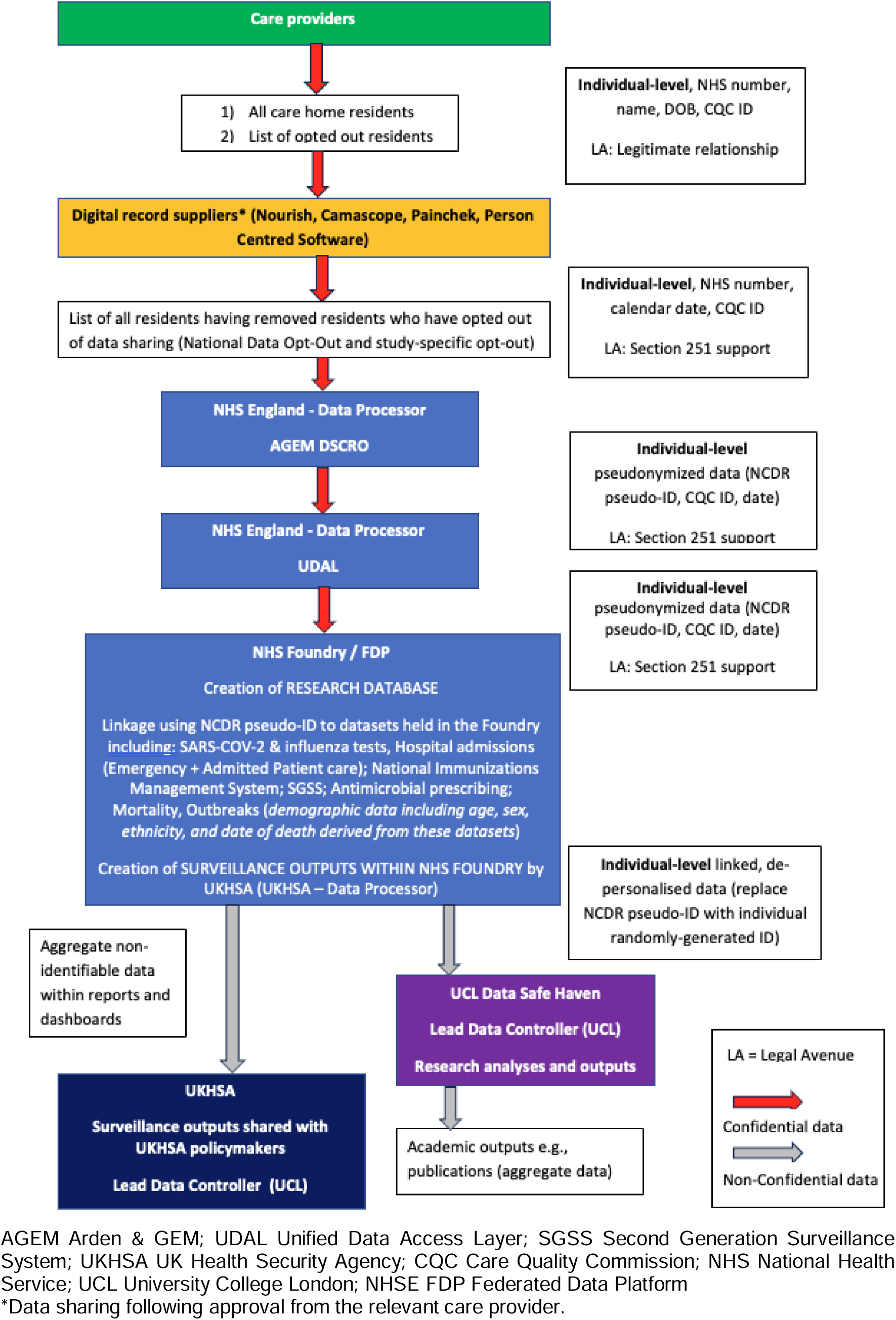
Data Flow Diagram. Diagram illustrating flow of data from providers to the study dataset. Red arrows represent confidential data and grey arrows demonstrate movement of non-confidential idea that has been pseudonymised / de-personalised. Research database will be created in secure data platform such as NHS Foundry or the Federated Data Platform (FDP).

**Table 1:** Data items and sources. Individual-level and facility-level data that will be included in study datasets alongside data sources.

| Data item | Data source / owner |
| --- | --- |
| **Individual-level** |  |
| - Care Home<br>- Date | Digital Social Care Record supplier |
| Vaccination (COVID-19, influenza)<br>- Type<br>- Date<br>- Dose number | National Immunisation Management System (NIMS), NHS England |
| Hospital admissions<br>- Date of admission<br>- Date of discharge<br>- Diagnostic codes for cause of admissions and other diagnoses | Admitted Patient Care (APC), NHS England |
| Emergency Department attendances<br>- Date of attendance<br>- Outcome (i.e., admission, transfer, discharge)<br>- Diagnostic codes | Emergency Care Dataset (ECDS), NHS England |
| Death<br>- Month of death<br>- Primary and secondary causes of death | Deaths, Office of National Statistics (ONS) |
| Results of virological / microbiological testing of clinical samples<br>- Date of test<br>- Organism detected<br>- Antimicrobial susceptibility (if applicable) | Second Generation Surveillance Systems (SGSS), UK Health Security Agency |
| Antimicrobial prescriptions issued in the community<br>- Prescription date | Antimicrobial prescriptions, NHS Business Services Authority |
| - Drug name<br>- Duration of course |  |

Pseudo-identifiers will be removed before the de-personalised data are transferred securely to UCL and stored in the UCL Data Safe Haven (DSH) (https://www.ucl.ac.uk/isd/services/file-storage-sharing/data-safe-haven-dsh). The DSH has been certified to the ISO27001 Information Security standard and conforms to NHS Digital’s Data Security and Protection Toolkit. It is built using a walled garden approach, where the data is stored, processed, and managed within the security of the system, avoiding the complexity of assured endpoint encryption. A file transfer mechanism enables information to be transferred into the walled garden simply and securely. Only named researchers within the research team have access to the research database within the DSH. All researchers seeking to use the DSH are required to complete regular training in information governance.

All residents will be given the opportunity to opt-out of data sharing by speaking to any member of care staff. They will also be able to notify the care home in writing that they do not wish to participate. The decision to opt out can also be made on behalf of a resident in accordance with their wishes, for example by their next of kin and/or a consultee, in line with the Mental Capacity Act 2005 and National Institute of Clinical Excellence (NICE) guidelines.^21,22^ Lists of individuals who have completed either the NHS opt out^23^ or the project specific opt out will be regularly updated and shared with DSCR suppliers who will remove these individuals from the data feed with NHSE. Information materials (posters, leaflets, information sheets, animation) will be distributed widely through participating care homes with details of how to opt-out.

#### Study Oversight

Oversight for the study is outlined in Figure 2. The Project *Steering Committee* will provide oversight of the study on behalf of the Sponsor (UCL) and Funder (UKHSA) and will be chaired by an experienced academic in social care policy who is not involved in study management. The *Stakeholder Oversight and Governance Group* will act on behalf of residents and families to oversee how data are used in the study and study outputs. This group will include representation from residents, relatives, staff, and providers and experts in information governance from care providers, UCL, UKHSA, and NHSE. It will meet two to three times per year. It will provide reassurance to care providers, residents, relatives, and staff that they maintain control of how their data will be used in the project and is a critical part of building trust between care home stakeholders and UKHSA to enable future studies and continued data sharing.

**Figure 2:**
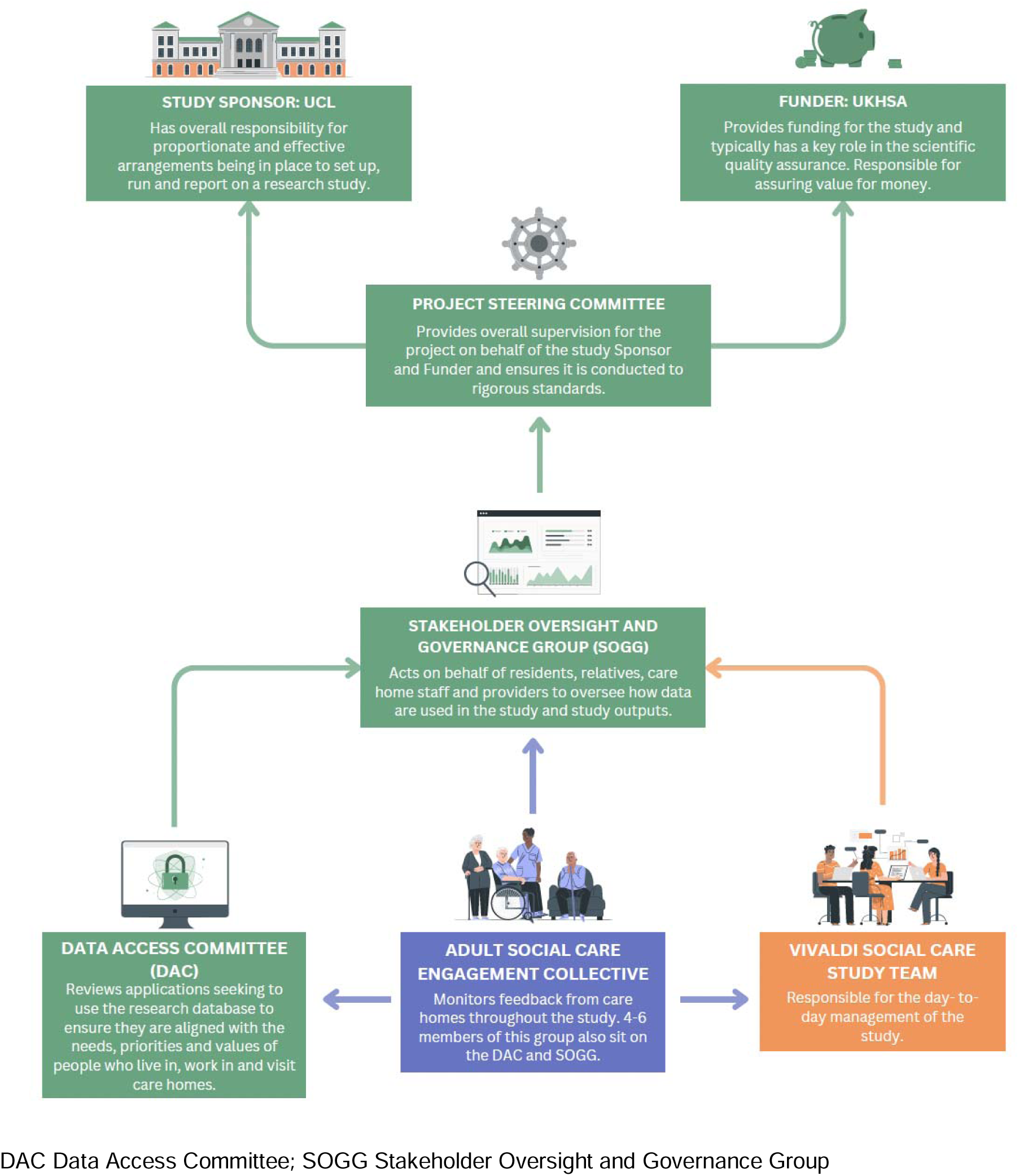
Study oversight diagram. Flow diagram representing the relationships between groups overseeing the project. Colouring of boxes and arrow demonstrates the role of each group: green represents oversight, orange is operational, and blue is advisory.

In parallel, a *Data Access Committee* will specifically oversee the use of the de-personalised, pseudonymised research database in observational / modelling research studies. This group will include representation from residents, relatives, staff, academics, and providers and will be responsible for reviewing and approving proposals from researchers who wish to use the linked dataset.

The *Adult Social Care Engagement Collective* will have an advisory role feeding directly into the oversight and operational groups, Figure 2. They will ensure the views of residents and relatives are represented throughout the project. This will consist of representatives from the care sector including providers, care managers and more junior staff; residents and their relatives and third sector organisations specialising in data and the rights of people accessing care.

#### Data Analysis & Statistical Plan

The sample size reflects the desire to generate findings that are generalisable across the care sector. In parallel it also acknowledges that there are considerable technical, logistical and governance challenges associated with setting up this study (collection of individual-level data from care homes at scale has not previously been attempted in England).

As there are approximately 11,000 care homes for older adults in England,^1^ if the target sample size (500-1500 care homes) is achieved, the sample would represent approximately 5-14% of all care homes. However, there is considerable diversity across the care sector, so it is very unlikely to be representative of all the different types of care homes. If the study is successful, and there is a desire to establish a long-term surveillance and linked data infrastructure in care homes, the intention is to work with the CQC and the UKHSA to understand which types of care homes are not included and to try and address these gaps by targeted recruitment. Given the complexity of the data flows and governance, the relatively short timescales, and the absence of a care home registry from which to sample, it is not feasible to develop a sampling frame and attempt to recruit a representative group of care homes in this pilot study.

As the goal of this project is to explore the feasibility of establishing sentinel surveillance and a research database enabling observational and future interventional studies, a Statistical Analysis Plan (SAP) has not been developed. All projects using the research database will be expected to develop a SAP before data analysis commences.

#### PPIE

This proposal has been informed by extensive engagement with care home residents, their relatives, care home staff, and providers including presentations at Care conferences and events held by the CQC, press releases, magazine articles, newsletters, webinars, and podcasts. Interactions have strongly informed the study design and specifically decisions to 1) seek section 251 support to access data without consent from residents and 2) focus the pilot study on care providers that use DSCR (due to the prohibitive workload associated with extracting information from paper-based systems).

Since October 2022, UCL, Care England and the OS have established two working groups (Engagement & Communications; Data Sharing & Information Governance) with representation from care providers, care home staff, and relatives of people who live in care homes. These groups considered how to engage and communicate with care homes, and issues related to data sharing and information governance.

The project team have visited care homes across England to discuss the project and the problem of infections in care homes. In addition, the study team have partnered with researchers from the DACHA study to develop VIVALDI Social Care ‘activity packs’, which are delivered by activity providers in participating care homes and provide a non-onerous mechanism to engage residents in the study and seek their views. A new collaboration has been established with the charity ‘Care Rights UK’ (https://www.carerightsuk.org/) who represent the views of residents and relatives. The study has been discussed with eight members of this organisation and they have also had input into the design of information leaflets and posters. A webinar for members of this organisation was also held to gather views regarding the use of data from residents without consent. The *Engagement & Communications workstream* are currently working with an animator to co-produce an animation to accessibly explain the concept of data sharing within the VIVALDI Social Care study.

The care home working groups, and Care Rights UK will continue to play a key role in the delivery of this project within the *Stakeholder Oversight & Governance Group*, *Engagement Collective*, and *Data Access Committee*. They will work with the research team to interpret results and advise on dissemination of findings to people who live and work in care homes.

Full details of our activities to date and those that will take place during the study are provided in the Appendix. Video recordings from engagement events are available to watch online: https://theoutstandingsociety.co.uk/case-study/vivaldi-care-home-study/.

#### Ethics

Ethical approval to set up and use the anonymised research database for observational / modelling studies on infection and AMR in care home residents has been provisionally obtained from the South-West - Frenchay Research Ethics Committee however all applications to use the database must first obtain approval from the study *Data Access Committee*. Use of the database in future interventional studies would require additional ethical approvals.

Provisional approval to collect data from residents without informed consent for the purposes of sentinel surveillance and research on AMR has been obtained from the Health Research Authority following review by the Confidentiality Advisory Group (CAG) under Section 251 of the NHS Act 2006. Two applications were submitted enabling use of the data for research (23/CAG/0134) and surveillance (23/CAG/0135).

#### Dissemination plans

Dissemination of outputs to key stakeholders will use the engagement workstreams outlined above. Results of analyses conducted using the VIVALDI Database will be published in peer-reviewed academic journals in a timely manner and posted on pre-print sites in cases where results are required to rapidly inform policy decisions. Results will also be presented to policymakers, academics, clinicians, and providers at national care conferences, academic infection conferences, and regular meetings with policymakers within UKHSA, Department of Health & Social Care (DHSC) & NHSE. Dashboards and surveillance reports will be accessible to key stakeholders including care providers and public health officials within UKHSA and policymakers in DHSC.

Study outcomes will be shared with participants in an end of study report. This will be developed in collaboration with the engagement workstream, to ensure that the results are clear and easy to understand for the care home population, and to determine the best format for this e.g., video. This will be published on the study website and this link will be widely disseminated so that family members and the wider care population are able to access it.

#### Conclusions

This project builds on the experience and lessons learned from the COVID-19 pandemic in care homes, which highlighted the urgent need for an effective data and research infrastructure to enable surveillance and research on infections and outbreaks and inform policy. This issue was also highlighted as a priority by the Chief Medical Officer in his Technical Report on the COVID-19 pandemic. Sentinel surveillance will improve our ability to measure the burden of infection and AMR in care homes and the research database will be made available to the researchers to facilitate observational and modelling studies. The database could also support future interventional studies that would be subject to additional ethical and governance approvals. The study will be delivered by a partnership comprising UCL, care sector organisations, policymakers, care providers, and digital suppliers. Data use and project delivery will be overseen by a stakeholder oversight group including relatives, residents, care home staff, and providers.

Although this project focuses on infectious diseases and AMR, if successful and there is continued support from residents and their relatives for data sharing, the database could be repurposed to enable a broader portfolio of care sector-led research studies addressing priorities such as falls or dementia.

## Data Availability

Data will be made accessible to researchers following approval of their project plan by the study’s *Data Access Committee* and confirmation that they have completed appropriate information governance training.

## Reporting Guidelines

University College London. Workflow: STROBE Checklist ‘VIVALDI Social Care Protocol’. https://doi.org/10.5522/04/24512893.v1.

Data are available under the terms of the Creative Commons “Attribution 4.0 International” Public License (CC BY 4.0 Public License).

## Author Contributions

LS, ZF, MG, CL, and MK conceptualised the study. LS, JC, GP, and GC acquired funding. ZF, BA, GP, GC, and CL administered the project. MK and LS wrote the first draft of the manuscript. All authors reviewed and edited subsequent drafts.

## Award Information

This work was supported by the National Institute for Health and Social Care Research Research Professorship (grant number 302435 to LS); the Wellcome Trust (grant number 222907/Z/21/Z to MK); Department of Health & Social Care (OS, AC, LS, KT); UK Health Data Research [grant number LOND1 to AH]; LS is funded by the NIHR University College London Hospitals Biomedical Research Centre. This work is independent research funded by the UK Health Security Agency. Views expressed in this publication are those of the authors and not necessarily those of the NHS, UKHSA or the Department of Health and Social Care.

## Acknowledgements

We thank those who have taken part in extensive engagement activities and provided their invaluable insights and feedback into the study design, particularly the two working groups (Engagement & Communications; Data Sharing & Information Governance). We also thank the staff and residents in care homes who participated in the VIVALDI study.

