## Appendix for "Protocol for VIVALDI Social Care: Pilot Study to reduce Infections, Outbreaks and Antimicrobial Resistance in Care Homes for Older Adults"

**Inventory of Engagement Work**

A detailed description of all our engagement activities that have informed the design of this study, with links to recordings of the meetings, is provided in the table below. Here we provide a high-level summary of our engagement activities to date:

Between September and December 2022, we held two task and finish groups to establish a plan to take VIVALDI Social Care forwards; this was followed by five working group discussions with 90 members of the care sector, including senior managers, academics, and policy-makers. These meetings aimed to develop a ‘high-level’ roadmap for the project and determine if it might be feasible. During this period, we also held three dissemination events with the wider sector, including friends and relatives of care home residents. These meetings focused on determining whether it is feasible to share data from residents for research purposes, approaches to consent, and how to overcome the technical challenges of data sharing (where the data will be held, how it will be extracted, who will act as data controllers, what project oversight is required and by whom). These meetings were led by the Outstanding Society (OS) and delivered in partnership with University College London (UCL) and Care England.

In January 2023, we established two separate task and finish groups: engagement & communications and data sharing & information governance.

### **Table 1: Task & Finish Group Membership**

| **Engagement & Communications Group** | **Data Sharing & Information Governance Group** |
| --- | --- |
| Director OS | Care England representative |
| Director OS | Provider |
| Policy Officer Care England | Provider |
| Professor of the Care of Older People | Policy representative |
| Care manager | Policy representative |
| IPC Specialist | Resident / relative representative |
| Senior Policy Officer | Resident / relative representative |
| Project Manager on the VIVALDI | Clinician |
| Co-Production Collective | Digital Software Provider |
| Relative | Local Authority director of Public Health |
| Healthwatch respresentative |  |
| Researcher in Residence - Provider |  |
| Head of Lifestyles - Provider |  |
| Senior manager |  |
| Relative |  |
| Senior Nurse |  |
| Senior Activity Coordinator |  |
| Student Nurse |  |
| Student Nurse |  |

OS Outstanding Society; IPC Infection Prevention & Control

The engagement workstream was responsible for considering the types of materials needed to explain the study, the level of detail required, and how to present this information in an accessible way. They were also tasked with reviewing the content of all participant-facing study materials, including plain language summaries, participant information sheets, and our video. Members of the group also participated in a podcast panel discussion for care home staff, relatives, and care home managers, which aimed to explain the study and our use of data to a lay audience.

The data governance group was tasked with considering consent models (informed consent versus an opt-out consent model), information governance (including how to simplify paperwork related to data sharing e.g. by providing template agreements and privacy statements), and oversight (who are data controllers and processors, what oversight mechanisms are required). The output of the data governance group was a

coproduced model for project governance and oversight with UCL, the OS and Care England acting as joint data controllers and an agreement to establish an oversight committee with representation from each of the three organisations, plus residents/relatives, policymakers, and academia. The oversight committee will be responsible for agreeing project outputs to ensure that all research is aligned with the needs and priorities of people who live and work in care homes.

In July and August 2023, we partnered with Care Rights UK to hold two resident/relative-focused events. At the first event we worked with 8 relatives to revise and improve the participant facing materials that our engagement group had produced. The second event was an online webinar (open to up to 1000 people) with a panel discussion aimed to give residents and relatives an opportunity to ask questions and raise their concerns.

In parallel, the research team have visited six care homes since November 2021 to talk to residents, relatives, and care home staff about the study. Plain language summaries for this study have also been reviewed by two members of the study-specific VIVALDI PPI group, who are relatives of care home residents.

The engagement and recruitment work package of our study is being led by Zoe Fry, one of the Directors of the OS who owned a nursing home for 13 years.

### **Table 2: Engagement activities undertaken for VIVALDI Social Care 2022-2023**

| **Description** | **Audience** | **Purpose** | **Further details & dates** |
| --- | --- | --- | --- |
| Online presentations to CQC Trade Association meetings | CQC & Trade Associations (who represent care providers) | To inform the CQC about the project  To raise awareness among Trade Associations ((TA’s)  For TAs to disseminate information to their members (care providers). Members to disseminate to providers relatives and residents. | March 2023  October 2022  March 2022 |
| Podcasts for the care sector, led by the Outstanding Society. These consist of short discussions, usually with a panel, to discuss specific aspects of the VIVALDI study in an accessible way) | Care providers, care home staff, relatives, stakeholders, the public | To raise awareness about the study To address specific concerns about the project, e.g. regarding the use of data and its storage | [Click here for podcasts](https://theoutstandingsociety.co.uk/case-study/vivaldi-care-home-study/resources/) |
| Attendance at Care Show including Panel Discussion entitle. Social Care Innovation.  Video recording of interview with Prof Martin Green (CEO of Care England, the largest care provider representative organisation) and Prof Shallcross, UCL about the VIVALDI study | Providers and Health and Social Care Professionals | Raise awareness of the project. Generate interest from care homes in taking part in a future study. Answer questions about the study. | October 2022  Interview Professor Martin Green and Professor Laura Shallcross  [Click Here for Care Show Interview](https://youtu.be/G9jrOBVkpxY?t=7517) |
| Care Show Showcase - panel discussion with relatives, providers, policy makers, UCL, DHSC Chief Nurse for Adult Social Care, Director of Nursing NIHR, CEO Nourish (software vendor), Digital Care Planning System | Providers and Health and Social Care Professionals | To raise awareness of the project To recruit care homes to take part To answer specific questions about the study | April 2023  [Click here to watch panel](https://www.youtube.com/watch?v=zD5xtv6fHis) [discussion Care Show 2023](https://www.youtube.com/watch?v=zD5xtv6fHis) |
| Care Show Showcase for VIVALDI Social Care – Onboarding | Providers and Health and Social Care Professionals | To onboard further care homes and update adult social care on the project.  To celebrate Care Rights UK involvement | October 2023 |
| Joint presentation from UCL and the Outstanding Society about VIVALDI Social Care to the Infection Prevention Society, Care home special interest group.  Virtual Meeting | Care home managers, community infection prevention leads (Usually nurses) | Raise awareness about the study among the infection prevention community  To answer specific questions /concerns about the study | 22nd February 2023 |
| Joint presentation from UCL and the Outstanding Society about VIVALDI Social Care to the Infection Prevention Society, Care home special interest group.  Face to Face meeting | Study day open to everyone who works with/in, or has an interest in Infection Prevention and Control and care homes | Raise awareness about the study among the infection prevention community.  To answer specific questions /concerns about the study | 24th May 2023 |
| VIVALDI Social Care Stakeholders Virtual Meeting.  Panel discussion with Deborah Sturdy, Ruth Endacott, the Directors of the OS and Professor Shallcross | 142 attendees, providers, residents, relatives, health and social care professionals | To update all stakeholders on the co-developed project roadmap and plans to pilot the study in 2023  To give stakeholders the opportunity to ask questions | 15^th^ December 2022  [Click here to view Stakeholders](https://theoutstandingsociety.co.uk/case-study/vivaldi-care-home-study/resources/#meeting) [Meeting](https://theoutstandingsociety.co.uk/case-study/vivaldi-care-home-study/resources/#meeting) |
| Stakeholders Virtual Meeting | 208 Attendees. providers, residents, relatives, health, and social care professionals | Progress update and the opportunity to ask questions ahead of the care show launch | March 2023  [Click here to veiw stakeholders](https://www.youtube.com/watch?v=NTqmNrqj6S8) [meeting](https://www.youtube.com/watch?v=NTqmNrqj6S8) |
| ARC Care Home Network Applied Research Collaboration Care Home Network Event NIHR | Researchers who work in social care and care providers and staff with an interest in research | Raise awareness about VIVALDI Social care and plans to pilot the study in 2023.  Identify opportunities for collaboration / draw on existing learning | 10th March 23  [Agenda ARC](https://arc-eoe.nihr.ac.uk/events/applied-research-collaboration-care-home-network-event) |
| Meetings with Individual providers |  | Discuss proposed plans and explore acceptability / willingness to get involved | Ad-hoc |
| Task and Finish Group | DHSC, Skills for care, providers, care associations, Digital social care, Care Associations | Two virtual meetings for a task and finish group to plan the future of VIVALDI | 2^nd^ September 2022  13^th^ September 2022 |
| Stakeholders engagement event | Video available for stakeholders unable to attend | To update a wider group of stakeholders on study progress to date and provide opportunities for questions. To form the working group | 27^th^ September 2022  [Click here for Stakeholders 27th](https://www.youtube.com/watch?v=X3-mrpyufsU) [Sept 22](https://www.youtube.com/watch?v=X3-mrpyufsU) |
| VIVALDI Social Care Working Group consists of 90 people.  WG1 1st working group meeting (ToR, introductions) |  | 1st working group meeting (ToR, introductions) | 18^th^ October 2022  [WG1 Click here to view](https://www.youtube.com/watch?v=EDIwiKHPWi4) |
| VIVALDI Social Care Working Group consists of 90 people.  WG2 Study outputs; ‘opt in’ versus ‘opt out’ consent |  | Focused discussion on potential outputs of the study and different approaches to obtaining consent for the use of data from residents (‘opt in’ versus ‘opt out’ consent) | 1^st^ November 2022  [WG2 Click here to view](https://www.youtube.com/watch?v=VsyMDiFCjuc) |
| VIVALDI Social Care Working Group consists of 90 people  WG3 How to extract data from care homes |  | Discussion of different approaches to extracting data (NHS numbers) for residents from care homes | 15^th^ November 2022  [WG3 Click here to view](https://www.youtube.com/watch?v=VPfRO4BH_V0) |
| VIVALDI Social Care Working Group consists of 90 people.  WG4 Data governance and oversight |  | Discussion regarding data governance and oversight in the project and what the study team need to put in place to make it easy for care homes to take part e.g. template agreements | 29^th^ November 2022  [WG4 Click here to veiw](https://www.youtube.com/watch?v=qWhLnRo_yO4) |
| VIVALDI Social Care Working Group consists of 90 people  WG5 Summary of progress to date and next steps |  | Summary of progress to date and our co-developed project roadmap. Overview of next steps and plans to pilot the study in 2023 | 13^th^ December 2022  [WG5 - Click here to view](https://www.youtube.com/watch?v=KkCz_QFz-l4) |
| Stakeholder engagement event | Update for residents and relatives. Providers to share the recording within the homes. | Engagement with residents and relatives | 6^th^ December 2022  [Stakeholders 6th Dec 2022 Click](https://www.youtube.com/watch?v=rRvO6TT_WDY) [here to view](https://www.youtube.com/watch?v=rRvO6TT_WDY) |
| Virtual Meetings January - March 2023  Engagement and Communication Workstream  January 25^th^ 15:30 – 16:30 Map existing models of engagement and communication for social care  February 16^th^ 15:00 – 16:00 Develop methodologies to improve engagement and communication strategies in the project.  March 7^th^ 10:00 – 11:00 Practicalities – what resources do we need for the engagement and communication strategy.  March 30^th^ 10:00 – 11:00 Agree the future aims of the engagement workstream | See Table 1 |  | [25th January 2023](https://www.youtube.com/watch?v=kv6_lpdCmNE) [16th February 2023](https://www.youtube.com/watch?v=kv6_lpdCmNE) [7th March 2023](https://www.youtube.com/watch?v=hTGvB0LnCD0) [30th March 2023](https://www.youtube.com/watch?v=NTqmNrqj6S8) |
| Virtual meetings January - March 2023  Governance and Oversight Workstream  January 26^th^ 12 – 1  History of the VIVALDI study, why we need research on infection and the importance of routine data in research.  February 16^th^ 11 - 12 Models for data sharing and oversight group including practicalities | See Table 1 |  | [26th January 2023](https://www.youtube.com/watch?v=qC2MOXmKOyE) [16th February 2023](https://www.youtube.com/watch?v=pEwEU6SN7WU) |
| DACHA | Residents via activity coordinators | To develop an activity pack to engage with residents in care homes to talk about research. | [Click here to find out more](https://dachastudy.com/#%3A~%3Atext%3DDACHA%20AIMS%2Cof%20innovation%20in%20care%20homes) [about DACHA.](https://dachastudy.com/#%3A~%3Atext%3DDACHA%20AIMS%2Cof%20innovation%20in%20care%20homes) |
| Project Discussion with Rights for Residents | Rights for residents’  founders (x3) | Initial discussion with founding members | 19^th^ June 2023  Rights for residents engaged with the project and fully supportive |
| Project Discussion Care Rights UK | Meeting with Director | To confirm if the merged Rights for residents and Care UK and fully supportive of the project | 29^th^ June 2023  Director agreed to be part of working group, support a meeting with members and be part of future steering group for the pilot |
| Care Homes Re-engagement | Providers | To confirm participation. To confirm digital supplier. To confirm if home would like a visit from the project team | [Teams engagement form -](https://forms.office.com/e/D5nvjdqkHR) [providers](https://forms.office.com/e/D5nvjdqkHR) |
| Engagement and Communication |  | Material Review Feedback and Discussion | 3^rd^ July 2023  [Engagement and Communication](https://www.youtube.com/watch?v=AwssE5giI7I) [3rd July](https://www.youtube.com/watch?v=AwssE5giI7I) |
| Working Group | UCL, Care England, OS, Care Rights UK | Material and Project Review | 10^th^ July 2023 |
| Care Home Visits | VIVALDI study team; Jenny Harries, Chief Executive UKHSA (Lansdowne visit), Lucy Chappell, Chief Scientific Advisory from the DHSC (Lansdowne visit), Michelle Dyson, Director General of Adult Social Care, (Lansdowne visit) | To learn about how care settings vary and to speak to residents, relatives and staff about their experiences and some of the challenges that they have faced in relation to the pandemic. To understand what matters to care home teams. The learnings from these visits will be applied to our current research studies, and will help to shape plans for our future research. This will ensure that our research is relevant and accessible to all members of the care community. | November 2023, Horkesley Manor Care Home; March 2023, Glentworth House Nursing Home;  January 2023, The Close Care Home;  November 2022, Ashdene Care Home;  May & September 2022, Lansdowne Care Home; November 2021, Eastbourne Care Home  [Details on each visit](https://www.ucl.ac.uk/health-informatics/research/vivaldi/vivaldi-patient-and-public-engagement) |
| Open Forum with Care Rights UK | Residents and relatives | Project Overview in partnership with Care Rights UK | 8^th^ August 2023  [Listen to recording](https://www.youtube.com/watch?v=sn-PpJDxsbg) |
| Development of VIVALDI animation with professional animator | VIVALDI team  Engagement & Communications Workstream | Developing animation to describe the study. Review of iterations of the animation | 29^th^ August 2023  [View the meeting](https://www.youtube.com/watch?v=OGcWNTwzKHU)  26^th^ September 2023 |
| **Communication & Engagement Workstream**  This working group aims to develop materials and innovative resources to support the pilot study and specifically to ensure that we are communicating key messages about the study to residents, families and care home staff clearly and concisely in ways that are accessible to them, The working group will also advise on the content of these resources and also the different types of approaches that can be used to engage with residents and families e.g. visiting care homes, online meetings, podcasts, conferences, videos, websites etc. | VIVALDI team  Engagement & Communications Workstream | Meeting 3 To agree on final documents/resources and communication strategy.  (Submission to CAG if required)  Meeting 2  To review draft materials and content  To review the draft communication strategy  Meeting 1  Agree on membership and ToR  To review existing materials and identify gaps for the pilot.  To identify modes of communication and frequency for all stakeholders | 3^rd^ October 23  26^th^ September 23  29^th^ Aug 23 |

ToR Terms of Reference; WG Working Group; CQC Care Quality Commission; OS Outstanding Society; UCL University College London; DACHA Developing resources and minimum dataset for Care Homes’ Adoption; ARC Applied Research Collaboration; NIHR National Institute of Health and Care Research

### **Table 3: Engagement events planned from November 2023 and during the 12-month pilot**

| **Description** | **Audience** | **Purpose** | **Details Links to resources and dates of events** |
| --- | --- | --- | --- |
| Open Meetings for Care Home Teams | Care Home Teams | Organise an open meeting for Care Homes to talk to them about the project. Same meeting once per week throughout September.  Record meetings and putting together one big FAQ document at the end to share the questions with all. | 30-minute drop-in sessions throughout September |
| Open Virtual meetings | Residents and Relatives | Inform and update | TBC Ongoing during 12-month pilot |
| **VIVALDI Social Care Adult Social Care Engagement Collective (ASCEC)** |  | The ASCEC will represent the views of residents, relatives, and providers throughout the project. We aim to establish a group of c.20 people, with the goal that at least five members are present at every meeting. This flexible approach will make it easier for people to participate because they are not expected to attend every meeting. The ASCEC will fulfil its role in three ways:   1) By facilitating feedback from providers to the trial team on the level of opt-outs, documentation, training and other feedback from residents, relatives and team members 2) By ensuring that ASCEC’s views are represented at the VIVALDI Stakeholder Oversight and Governance Group 3) By ensuring that ASCEC’s views are represented at meetings of the VIVALDI Data Access Committee | 1^st^ meeting 14^th^ November 2023 then monthly thereafter |
| **VIVALDI Social Care leads Training** | Leads | Coproduced training by OS, UCL, Skills for Care, NIHR | 27^th^ November  Then January 2024 |
| **Care Show April 2024** | Adult Social care | Update on the project | April 2024 |
| **QNI** QNI IPC Champions Network, this is funded by DHSC for staff working in Adult Social Care. We currently have approx. 1600 members | IPC Champions | Overview of VIVALDI Social Care | January 2024 |
| **October 2024 – Care Show Birmingham** | Adult Social care | Update on the project | Oct 2024 |
| **Launch of the webpage** | All | Inform  Resources | Nov 23 |

### **Table 4: Documents developing to engage with stakeholders**

| **Description** | **Audience** | **Purpose** |
| --- | --- | --- |
| Activity packs (refine/develop existing tools) | Residents | To use by activity coordinators |
| Podcasts | Providers / Residents / Relatives / Health and Social Care Professionals | To inform and update |
| Residents, relatives, and Team Meetings | Residents, relatives, and team meetings | ‘Home VIVALDI Lead’ who will be taught at an away day will disseminate information |
| Display Boards | Visitors to care home | Visual to signpost |
| Social media | All | Raise the profile and inform |
| News releases | CQC, DHSC, NIHR |  |
| Video (recorded at care show) | Providers, Nurses, policymakers, academics | Explain what Vivaldi social care is. Recruit care providers to take part in the study.  Raise profile of the project |
| Explainer video (3-5 mins) | Relatives, care home staff, residents | Explain what Vivaldi social care is. Explain why we need to use data from residents |
| Poster | Residents and relatives | Raise awareness of the study Tells residents and family members how to opt out of data sharing |
| Brochures | Residents  Relatives  Staff | More detailed explanation about the study  Signpost to more information about data sharing etc |
| Website | All | Source of detailed information and FAQs about the study Resources for providers History of the VIVALDI |
| Activity packs | Residents | Mechanism to engage a subset of residents in the project and get their views |
| Boiler plate text for providers | Care providers (to share with residents and families). Can be added to providers websites and given to residents/ families when they enter the care home | Short summary of what the study involves and how data will be used + draft text for privacy notice |
| Conference presentations | Various |  |
| Glossary | All | Explain abbreviations and organisations |
| FAQ | Providers, residents, relatives |  |
| In depth overview of VIVALDI Social Care | Providers, residents, relatives, health and social care professionals | This will be available on the webpage for anyone wanting to know the detail behind the project – all explainer documents will signpost to the details |
